# Improvement in Kidney Function in Patients with Chronic Hepatitis B and Chronic Kidney Disease after switching to Tenofovir Alafenamide Fumarate: A Systematic Review with Single Arm Meta-analysis

**DOI:** 10.1101/2025.06.29.25330521

**Authors:** João Galdino de Pascoa Junior, Ramon Huntermann, Victor Machado Viana Gomes, Frederico de Sousa Marinho Mendes Filho, João Marcelo Vallim Bertozzi, Paulo Ricardo Gessolo Lins

## Abstract

**Objective:** Tenofovir Disoproxil Fumarate (TDF) is effective in treating Hepatitis B Virus (HBV) but has been associated with nephrotoxicity. In contrast, Tenofovir Alafenamide Fumarate (TAF) has emerged as a safer alternative, reducing kidney exposure while maintaining antiviral efficacy. This meta-analysis evaluates improvements in kidney function following the switch from TDF to TAF.

**Methods:** Our study was registered in PROSPERO (CRD42024565358) and included 10 randomized controlled trials (RCTs) involving 1,179 patients with Chronic Kidney Disease (CKD). We compared renal function before and after switching to TAF

**Results:** Significant improvements in glomerular filtration rate (GFR) were observed, indicating enhanced kidney function post-switch. The findings confirm that TAF demonstrates a superior renal safety profile compared to TDF, particularly in long-term treatments.

**Conclusion:** The clinical relevance of TAF for HBV patients with CKD aligns with current guideline shifts favoring TAF. Despite limitations such as high heterogeneity, this study supports TAF as a safer management strategy for HBV patients with CKD, demonstrating improved kidney outcomes and reduced nephrotoxicity risks. These findings support its broader use in clinical practice and highlight the need for further research on long-term renal outcomes.

## INTRODUCTION

Chronic HBV remains a significant global health challenge, affecting millions and leading to severe liver complications such as cirrhosis and hepatocellular carcinoma^1,2^ and effective long-term antiviral therapy is essential to suppress viral replication and reduce disease progression^1,3^.

The current first-line treatment for chronic HBV includes nucleoside/nucleotide analogues, specifically Entecavir, TDF, and TAF, as well as (Peg-IFN)^3^. These agents are recommended for their efficacy in suppressing HBV replication, reducing liver inflammation, and lowering the risk of cirrhosis and hepatocellular carcinoma^1,3^. Nucleoside/nucleotide analogues are particularly favored due to their oral administration and favorable resistance profiles^3,4^.

Tenofovir disoproxil fumarate (TDF) has been widely adopted for its potent antiviral efficacy^5^. However, its long-term use has raised concerns about renal safety, especially in patients with pre-existing kidney disease or those requiring prolonged therapy^1,4^. TDF has been linked to nephrotoxicity, including decreased glomerular filtration rate (GFR), proteinuria, and Fanconi syndrome. These effects are primarily attributed to the accumulation of tenofovir within proximal tubular cells, where it induces mitochondrial dysfunction by depleting mitochondrial DNA, disrupting oxidative phosphorylation, and impairing cellular energy metabolism. This mitochondrial toxicity can ultimately lead to acute tubular injury and chronic kidney disease, particularly in patients with underlining CKD^6,7^.

To overcome the renal and bone safety concerns associated with TDF, tenofovir alafenamide (TAF) was developed as a novel prodrug. Like TDF, TAF inhibits HBV reverse transcriptase, but its improved stability in plasma and selective intracellular activation result in higher hepatocyte delivery at lower doses^4,8^. This pharmacokinetic advantage maintains potent antiviral activity while significantly reducing systemic exposure and minimizing systemic toxicity (mainly renal and bone toxicities)^9^.

Recent evidence reinforces the improved safety profile of TAF. A study by Sax et al. demonstrated that switching from TDF to TAF improves renal function markers, such as reductions in proteinuria, increases in estimated glomerular filtration rate (eGFR), and lower levels of tubular injury biomarkers^10^. These findings are further supported by a large real-world cohort study by Farag et al., which followed chronic hepatitis B patients for up to 160 weeks after switching to TAF. The study showed that in patients previously treated with TDF, the decline in eGFR was halted or even reversed after switching to TAF, particularly in those with moderate-to-advanced CKD (eGFR <60 mL/min). Notably, improvements were also observed in patients with stage 2 CKD (eGFR 60–89 mL/min), suggesting that the renal benefits of TAF may extend beyond currently recommended thresholds^11^. These consistent findings across clinical trial and real-world settings strengthen the rationale for transitioning at-risk patients to TAF to mitigate renal complications during long-term antiviral therapy.

In this context, the present single-arm meta-analysis aims to synthesize data from the past decade to evaluate renal outcomes in patients with chronic HBV and CKD who switched from TDF to TAF. By focusing on this high-risk population, the study seeks to determine whether TAF provides a safer, yet equally effective, antiviral strategy for long-term HBV management.

## METHODS

This systematic review with meta-analysis was registered in the International Prospective Register of Systematic Reviews (PROSPERO) under protocol CRD42024565358. This study was designed following the Preferred Reporting Items for Systematic Reviews and Meta-analyses (PRISMA) reporting guidelines.

### Study eligibility

We included studies meeting the following criteria: (1) RCTs or cohort studies evaluating renal function before and after switching to TAF in patients with CKD. We excluded studies with (1) incomplete data, (2) kidney and/or liver transplant, and (3) overlapping patient populations. No restrictions on publication date or language were applied.

### Search strategy and data extraction

MEDLINE, Cochrane, and Embase databases were systematically searched on July 01, 2024. The search strategy was as follows: (“chronic hepatitis B”) AND (“chronic kidney disease”) AND (“tenofovir alafenamide” OR “TAF”) AND (“tenofovir disoproxil fumarate” OR “TDF” OR “Adefovir” OR “ADV”). We extracted data for (1) the number of patients with CKD prior to the change to TAF and (2) a number of patients with an increase of, at least, one stage of CKD. All identified articles were systematically assessed using the inclusion and exclusion criteria. Article selection and data extraction were undertaken independently by at least two reviewers. Emails were sent to the authors in case data was not available in the study text. The data on the number of patients with increased eGFR (main outcome) was extracted by reading the text and arranging it in Excel spreadsheets. Disagreements were resolved by consensus.

### Quality assessment

The quality assessment for this meta-analysis followed a rigorous and standardized approach to ensure the reliability and validity of the included studies. The risk of bias assessment was conducted using the Cochrane Risk of Bias (RoB2) tool to evaluate RCTs^12^. Additionally, a Funnel Plot and Egger’s Test were performed^13^. Each study was independently assessed by two reviewers for potential biases, including selection bias, performance bias, detection bias, and reporting bias. Discrepancies were resolved through discussion or by consulting a third reviewer when necessary. Studies were rated based on methodological quality, including the adequacy of randomization, blinding, and completeness of outcome data.

### Sensitivity analysis

For the sensitivity analysis, several tests were conducted to assess the robustness of our findings and to determine whether the results were influenced by specific studies or methodological assumptions. First, a leave-one-out analysis was performed, in which each study was sequentially excluded to evaluate its impact on the overall effect size. This approach helped identify whether any single study had a disproportionate influence on the meta-analysis outcomes. Additionally, a meta-regression was conducted based on available baseline characteristics—namely age, eGFR, and sex—to explore potential sources of heterogeneity. The influence of studies deemed to have a high risk of bias was also examined by performing a separate analysis excluding these studies.

### Data analysis

A comprehensive approach was employed to synthesize and interpret the findings from the studies included. Statistical analyses were performed using a random-effects model to account for potential heterogeneity between studies^13,14^. Treatment effects for binary endpoints and mean differences (MD) for continuous variables were calculated, both with 95% confidence intervals (CIs). The Mantel–Haenszel test was applied to all binary endpoints.

Heterogeneity across studies was assessed using the I^2^ statistic, with values of 25%, 50%, and 75% interpreted as low, moderate, and high heterogeneity, respectively. In cases of substantial heterogeneity (I^2^ > 50%), subgroup analyses were conducted to investigate potential sources of variation, including differences in patient characteristics, study design, or follow-up duration. Publication bias was assessed through visual inspection of funnel plots and quantitatively evaluated using Egger’s regression test; asymmetry in the funnel plot was considered indicative of potential publication bias. Statistical significance was set at a two-tailed p-value < 0.05. All statistical analyses were conducted using R software (version 2024.12.1 build 563), employing the packages meta^15^, metafor^16^, and dmetar^17^ for meta-analysis, meta-regression, and bias assessment, ensuring a rigorous and reproducible synthesis of data.

## RESULTS

Our systematic search initially identified 69 potential articles, as illustrated in Figure 1. After removing duplicates and excluding studies that did not meet the inclusion criteria based on title and abstract screening, 13 articles remained for full-text review. Of these, 10 RCTs met all eligibility criteria and were included in the meta-analysis. One was excluded for being a study protocol, and two others lacked sufficient outcome data. In total, the meta-analysis included data from 1,179 patients with CKD who switched from TDF to TAF.

**Figure 1.**
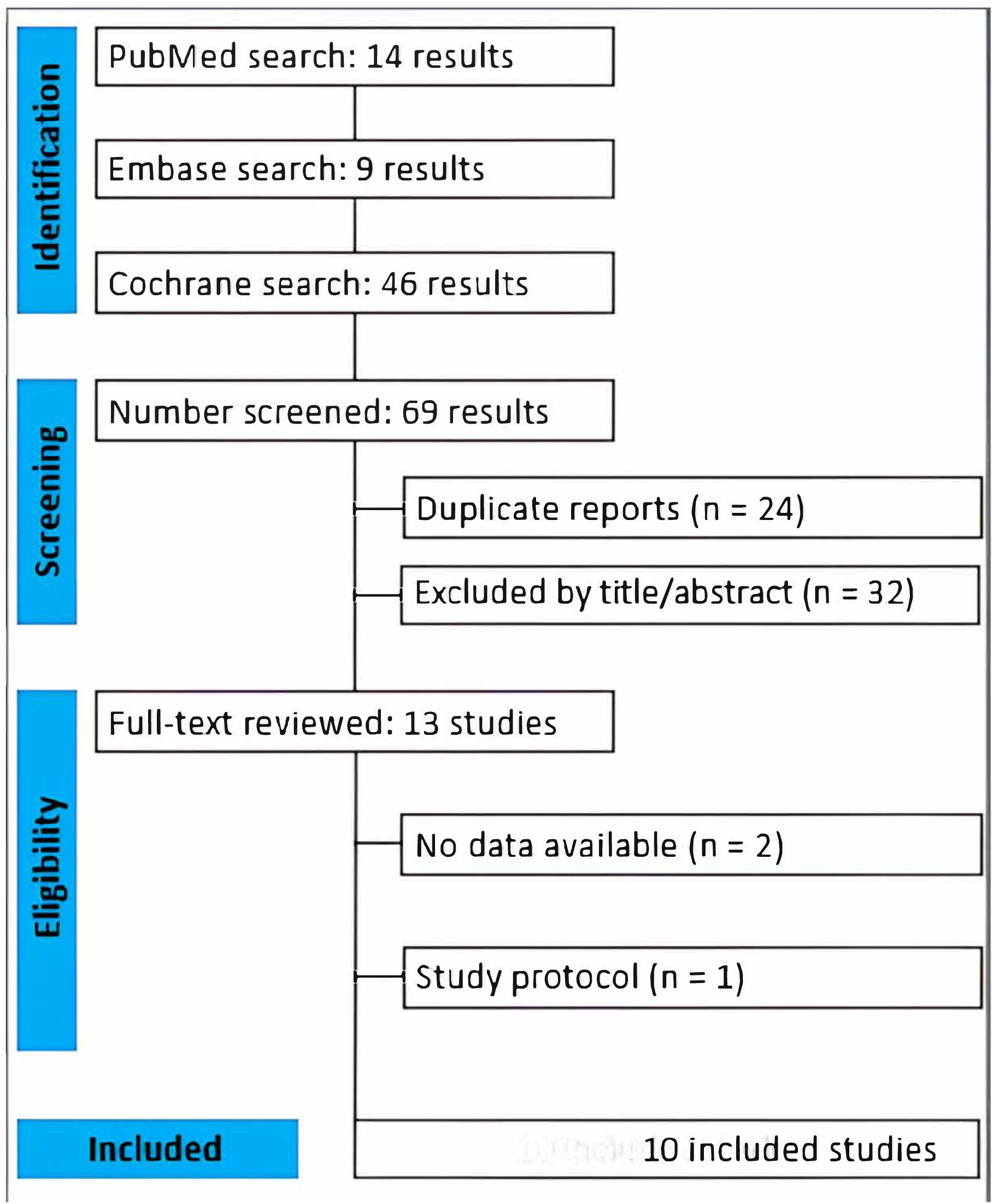
PRISMA flow diagram of studies selection.

Risk of bias was assessed using the RoB 2 tool (Figure 2). Publication bias was evaluated through a funnel plot (Figure 3) and Egger’s test, which did not indicate significant asymmetry (intercept = -1.38; 95% CI: -4.40 to 1.63; p = 0.39), suggesting a low likelihood of publication bias.

**Figure 2.**
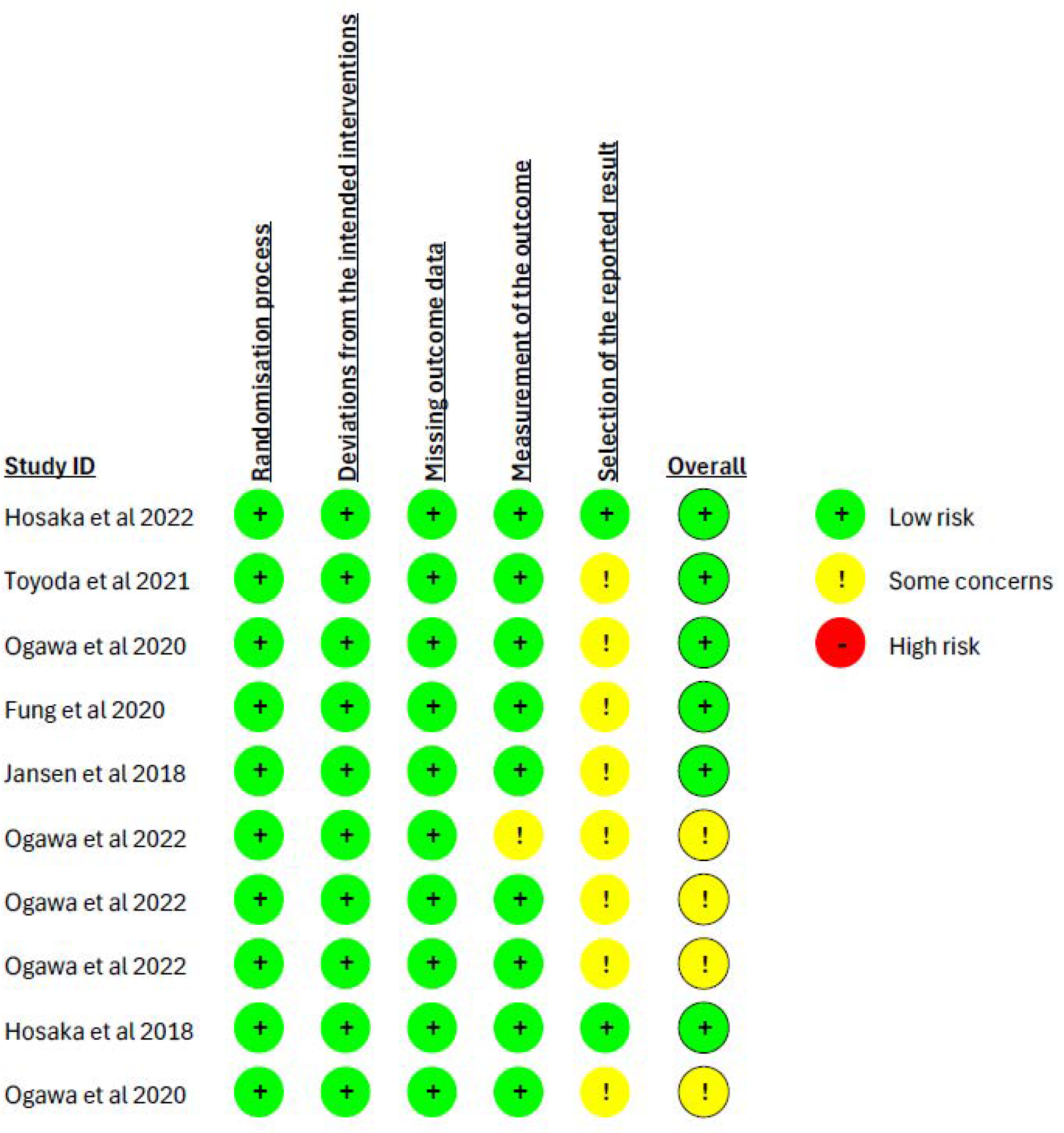
Cochrane Risk of Bias (RoB2)

**Figure 3.**
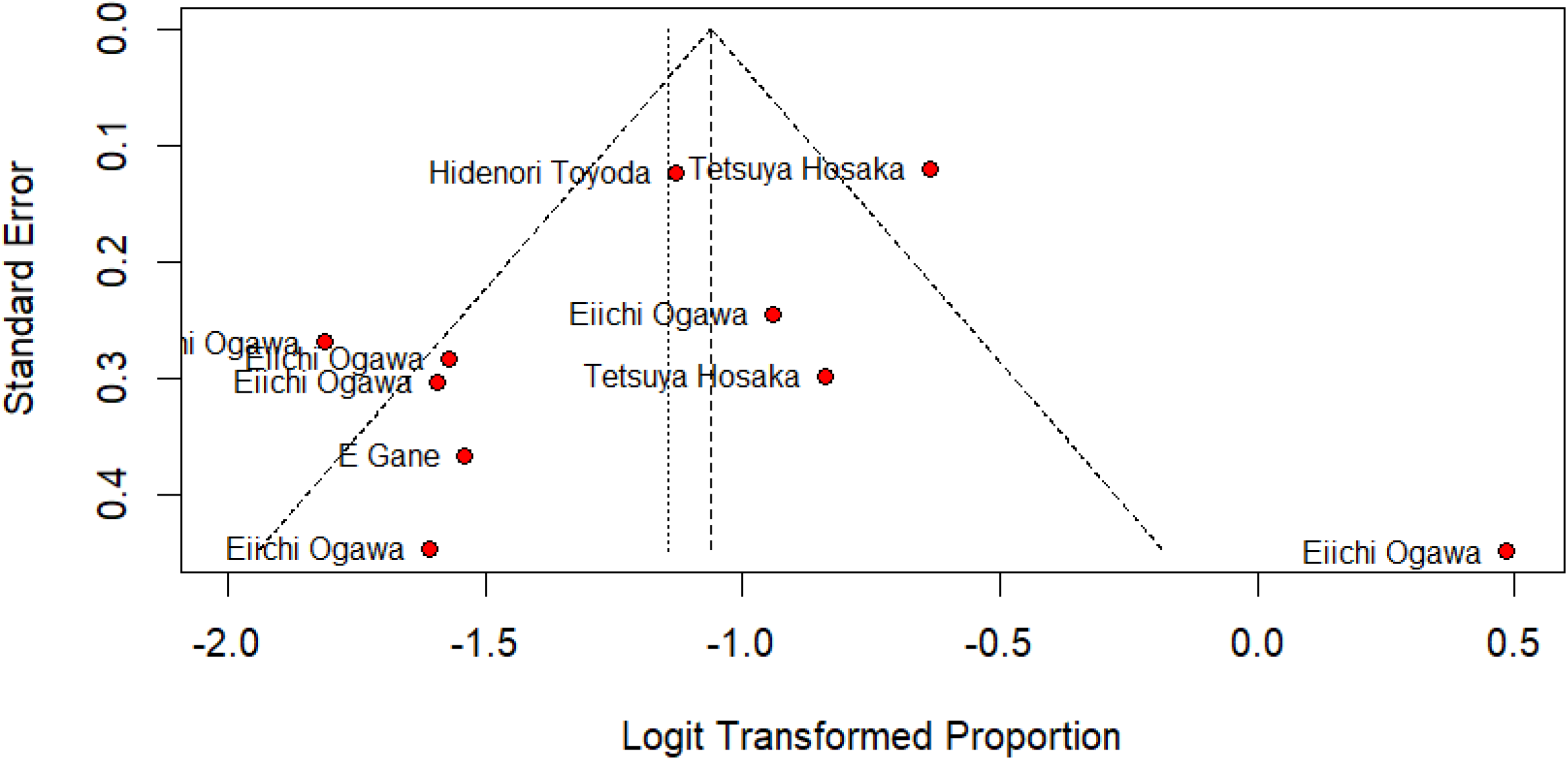
Funnel Plot.

The meta-analysis focused on evaluating the impact of switching from TDF to TAF on kidney function in patients with HBV and CKD. Although TDF is effective in suppressing HBV, its long-term use has been associated with nephrotoxicity, including reductions in GFR and other renal complications. TAF, a newer prodrug of tenofovir, provides equivalent antiviral efficacy with significantly lower systemic exposure, potentially offering improved renal outcomes.

The pooled analysis demonstrated a significant improvement in GFR following the switch to TAF. Among the 1,179 patients included the OR for GFR improvement was 24.13 (95% CI: 18.31–31.09; p < 0.01), indicating a statistically significant benefit. Despite high heterogeneity (I^2^ = 79%), the direction and magnitude of the effect were consistent across studies, supporting the renal safety profile of TAF compared to TDF (Figure 4). Detailed data used for the analysis are provided in the supplementary material.

**Figure 4.**
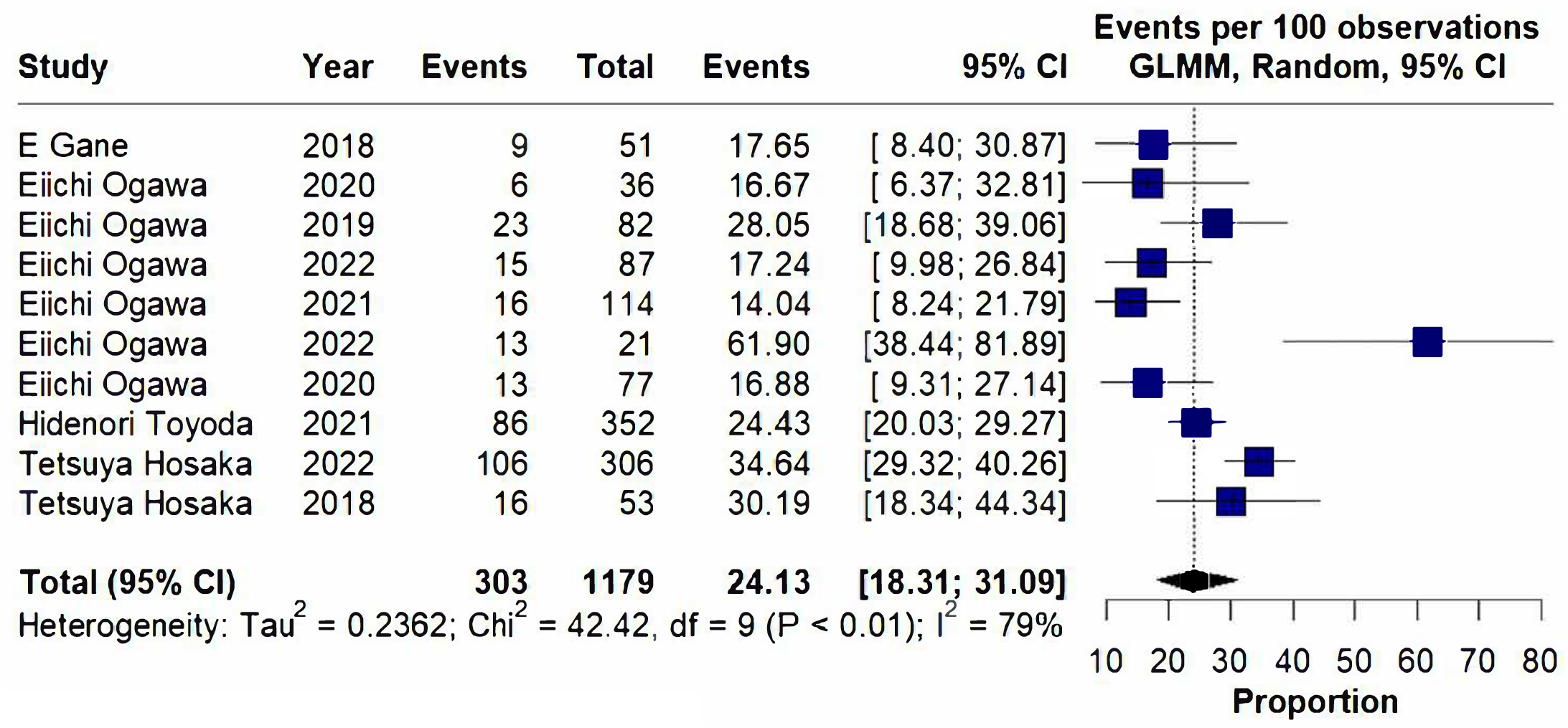
Forest plot graphic.

It is important to note that no secondary outcomes—such as progression to ESRD, markers of tubular injury, or bone health indicators—were consistently reported across studies. As such, this meta-analysis was limited to evaluating changes in GFR.

High heterogeneity was observed in four of the included studies—Hosaka et al.^18,19^, Fung et al.^11^, and Ogawa et al.^20^—which can be attributed to several factors. These include the fact that eGFR was not the primary outcome in some studies, small sample sizes, wide age variability, diverse ethnic backgrounds, the presence of comorbidities beyond CKD, and prior exposure to combination antiviral therapies before switching from TDF to TAF. Despite these sources of variability, the baseline characteristics of the study populations remained broadly aligned with our inclusion criteria, supporting the internal consistency of the analysis. According to the GRADE framework, this alignment reinforces the overall robustness of the findings.

To further explore the impact of heterogeneity, sensitivity analyses were performed. Excluding the single most heterogeneous study yielded an OR for eGFR improvement of 22.29 (95% CI: 17.75–27.61; p < 0.01), maintaining statistical significance (Figure 5). When all four highly heterogeneous studies were excluded, the OR adjusted to 18.70 (95% CI: 14.77–23.40; p = 0.16), indicating reduced precision but still suggesting a favorable trend toward renal benefit with TAF.

**Figure 5.**
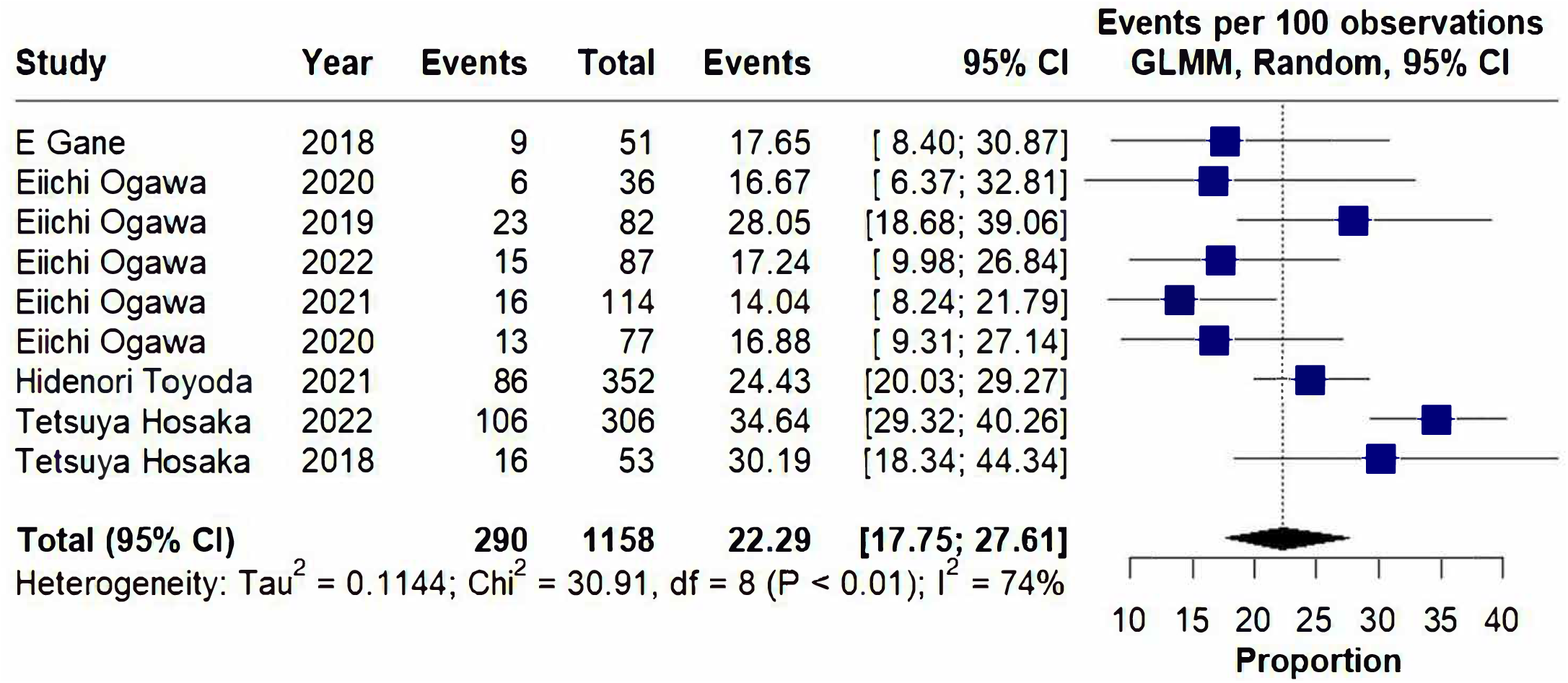
Forest plot graphic without the most heterogenous.

In addition, a mixed-effects meta-regression was conducted to investigate whether baseline variables could explain part of the observed heterogeneity. Significant associations were found with baseline eGFR (τ^2^ = 0.15, τ = 0.39, intercept = -1.35; p < 0.001) and age (τ^2^ = 0.07, τ = 0.28, intercept = -1.80; p < 0.001), while no significant influence was detected for male (p = 0.80) or female sex (p = 0.77). These results suggest that additional, unmeasured variables may be contributing to inter-study variability.

Beyond statistical significance, the findings underscore the clinical relevance of switching from TDF to TAF in patients with HBV and CKD. Given the established nephrotoxicity risk associated with prolonged TDF use, transitioning to TAF not only improves renal function markers such as eGFR but may also reduce long-term renal damage. These results strengthen current clinical preferences favoring TAF, particularly for patients with pre-existing renal impairment or those at risk of kidney dysfunction due to long-term antiviral therapy. Overall, the evidence supports prioritizing TAF as the preferred therapeutic option in the management of HBV in patients with compromised renal function.

## DISCUSSION

This systematic review and meta-analysis evaluated renal outcomes in 1,179 patients with CKD and chronic HBV infection who transitioned from TDF to TAF across 10 randomized controlled trials. The primary finding was a significant improvement in eGFR following the switch, with a substantial proportion of patients experiencing an improvement of at least one CKD stage. These results provide robust support for the renal safety of TAF and reinforce current clinical preferences for its use in managing HBV in patients with renal impairment.

Although TDF is effective in viral suppression, its long-term use has been consistently linked to nephrotoxicity. In contrast, TAF delivers the active metabolite more efficiently at lower systemic concentrations, reducing renal exposure and associated toxicity. Our findings reinforce this pharmacological rationale, supporting the preference for TAF in patients with existing renal dysfunction or those requiring prolonged antiviral therapy^1,2^.

Importantly, the sensitivity and meta-regression analyses demonstrated that the observed renal benefits of TAF remained consistent despite high heterogeneity among studies. This heterogeneity, observed in studies such as Hosaka et al.^18,19^, Fung et al.^11^, and Ogawa et al.^20^, was attributed to differences in study design, baseline characteristics, comorbidities, and prior treatment regimens. Nevertheless, the methodological consistency and comparable patient profiles across studies allowed us to maintain confidence in the pooled effect estimates. Even when the most heterogeneous studies were excluded, the renal benefit of TAF persisted, highlighting the robustness of the findings.

Despite the strength of the evidence, this meta-analysis also revealed important limitations in the current literature. Notably, secondary outcomes such as progression to ESRD, markers of tubular dysfunction, or bone health parameters—particularly relevant given the known impact of nucleotide analogues on bone mineral density—were inconsistently reported or entirely absent. This gap limits a more comprehensive assessment of TAF’s full safety profile and underscores the need for future studies to incorporate broader renal and skeletal endpoints.

Importantly, the World Health Organization (WHO) has reaffirmed the global goal of eliminating hepatitis B as a public health threat by 2030^21^. However, progress has been slow, with fewer than 3% of the estimated 254 million people living with chronic HBV currently receiving treatment^1,2,21^. In this context, strategies that prioritize not only antiviral efficacy but also safety and long-term tolerability—such as expanding access to TAF—are essential^1^.

The updated WHO 2024 guidelines emphasize simplifying treatment algorithms and expanding treatment eligibility, especially in LMICs where complex diagnostics and long-term monitoring are often not feasible. Integrating renal safety considerations into these simplified frameworks can help optimize treatment outcomes and prevent avoidable complications^21^. Policymakers should consider including renal outcomes in cost-effectiveness analyses and national treatment protocols to support broader access to TAF, ultimately contributing to the 2030 elimination agenda.

This study also aligns with the scientific community’s efforts to prioritize drug safety alongside efficacy. Given the well-documented nephrotoxicity associated with long-term TDF use^9^, this meta-analysis consolidates evidence of TAF’s superior renal safety profile, reinforcing its inclusion in clinical guidelines for hepatitis B management. These findings not only validate the ongoing shift toward TAF in clinical practice but also support its broader use among patients with pre-existing renal impairment. Additionally, this meta-analysis strengthens the evidence base by synthesizing data from diverse studies, addressing gaps in real-world data, and helping healthcare providers make evidence-based treatment decisions. By contributing to the growing body of research on reducing drug-induced organ toxicity, these findings may also drive further innovations in antiviral therapies and public health strategies.

In HIV-1 treatment, regimens containing TAF have already demonstrated non-inferiority in antiviral efficacy while offering improved safety regarding renal, bone, and lipid metabolism outcomes. In a meta-analysis by Tao *et al*., an OR of 0.31 (95% CI: 0.18–0.55) indicated a significantly lower risk of renal events when comparing TAF to TDF^22^. Conversely, another meta-analysis published the same year by Pilkinton et al. found no significant difference in renal adverse event rates between groups receiving HIV regimens containing either TAF or TDF. However, in that study, the number of reported renal events was relatively small (only 3 out of 4,665 patients in the TAF group and 6 out of 4,227 in the TDF group), potentially limiting its conclusions^23^.

This study has several limitations. Most notably, it lacks a control group directly comparing continued TDF use with the switch to TAF, which limits the ability to draw definitive comparative conclusions. Additionally, the analysis included only patients with CKD and chronic HBV infection, which may restrict the generalizability of the findings to broader HBV populations and contribute to the observed heterogeneity. The high heterogeneity across studies likely reflects variability in baseline patient characteristics, study designs, comorbidities, and follow-up durations. Moreover, not all studies provided complete data on eGFR changes, which may have influenced the precision of the pooled estimates.

Despite these limitations, the findings offer important insights into the renal safety profile of TAF and underscore its potential as a safer alternative to TDF, particularly for patients with compromised renal function. The consistency of the observed eGFR improvements supports the clinical relevance of transitioning to TAF in this vulnerable population.

Furthermore, the present study did not evaluate other clinically significant outcomes related to CKD and nephrotoxicity. While eGFR is a central marker of renal function, it does not capture the full spectrum of renal injury. Relevant endpoints such as progression to end-stage renal disease (ESRD), presence of tubulopathy, phosphaturia, alterations in bone metabolism, and serum phosphorus disturbances were not reported consistently in the included studies and therefore could not be analyzed. Future RCTs with broader outcome measures, inclusion of control groups, and more diverse patient populations are needed to provide a more comprehensive assessment of the renal and metabolic safety of long-term antiviral therapy—particularly in the context of tenofovir-associated tubular toxicity.

## CONCLUSION

This study provides compelling evidence supporting the renal safety advantage of TAF over TDF in patients with CKD and chronic hepatitis B. Although limited by the absence of a direct control group and the lack of data on additional renal endpoints— such as progression to ESRD and markers of tubular dysfunction - the consistent improvement in eGFR across studies suggests that TAF is a safer alternative for patients at increased risk of nephrotoxicity.

These findings underscore the clinical value of incorporating TAF into treatment strategies for vulnerable populations and support its broader adoption in HBV management guidelines, particularly in settings where long-term renal preservation is critical. Future research should aim to include more diverse patient populations and assess a broader range of renal and metabolic outcomes to further validate and expand upon these results. Such evidence will be essential for informing precision-based, kidney-conscious antiviral strategies in the ongoing effort to optimize HBV care.

## Abbreviations

CKD: Chronic Kidney Disease
eGFR: Estimated Glomerular Filtration Rate
ESRD: End-stage renal disease
GFR: Glomerular Filtration Rate
HBV: Hepatitis B virus
Peg-IFN: PegInterferon
RCT: Randomized clinical trial
TAF: Tenofovir alafenamide
TDF: Tenofovir disoproxil fumarate

## DECLARATION

### Authors’ Contributions

Pascoa Junior conceived of the presented idea. Pascoa Junior and Huntermann developed the theory and performed the computations. Gomes and Mendes Filho verified the analytical methods. Bertozzi and Lins supervised the findings of this work. All authors discussed the results and contributed to the final manuscript.

### Data Availability

The data that supports the findings of this study are available in the supporting information of this article.

